# IL-6 Receptor Antagonists and Severe Post-COVID-19 Outcomes: An Emulated Target Trial

**DOI:** 10.64898/2026.02.27.26347274

**Authors:** Zachary Butzin-Dozier, Manav Kumar, Yunwen Ji, Lin-Chiun Wang, A. Jerrod Anzalone, Eric Hurwitz, Rena C. Patel, Rachel Wong, Carolyn Bramante, Benjamin Sines, the National Clinical Cohort Collaborative (N3C) Consortium

## Abstract

**Background:** Interleukin-6 (IL-6) is a cytokine that plays a key role in systemic hyperinflammation and may mediate the relationship between acute COVID-19 and severe long-term outcomes such as Long COVID or death. IL-6 modulating drugs may reduce patients’ risk of severe post-COVID-19 outcomes.

**Methods:** We conducted an emulated target trial in a retrospective cohort of patients with moderate-to-severe rheumatoid arthritis who were prescribed IL-6 receptor antagonists (sarilumab or tocilizumab, pooled treatment) or other biologic agents (anakinra or baricitinib, pooled comparator) in 2022. We compared the 12–month cumulative incidence of mortality and Long COVID (diagnosed and probable) between groups using Super Learner and targeted maximum likelihood estimation, adjusting for covariates of interest.

**Results:** In our cohort of 3,553 patients, we found that prescription of IL-6 receptor antagonists was associated with a lower 12-month cumulative mortality (adjusted relative risk (aRR) 0.40, 95% CI 0.27, 0.59), diagnosed Long COVID aRR 0.42, 95% CI 0.23, 0.78), and probable Long COVID (aRR 0.71, 95% CI 0.61, 0.83), compared to prescription of other biologic agents, among rheumatoid arthritis patients.

**Conclusions:** IL-6 receptor antagonists may prevent the incidence of severe post-COVID-19 outcomes, such as Long COVID or mortality. This supports the hypothesis that IL-6 may be a mechanistic biomarker of COVID-19 sequelae and that acute COVID-19 severity may mediate this relationship.

---

Patients with prolonged symptoms of COVID-19 have persistent inflammatory dysregulation, and both Long COVID and acute COVID-19 are associated with dysregulation of cytokines, particularly interleukin (IL)-6. ^1–5^ Community cohort studies have demonstrated elevated IL-6 levels in patients with Long COVID compared with those who have fully resolved symptoms after acute SARS-CoV-2 infection.^5,6^ Furthermore, elevated IL-6 levels during inpatient treatment of acute COVID-19 are associated with a twofold increase in the risk of Long COVID compared with normal IL-6 levels at the time of acute infection.^4^

Investigators have proposed mechanistic causal models that support IL-6 as a mediator of the relationship between acute infection and Long COVID.^1–5,7–10^ IL-6 may mediate long-term neurological symptoms of COVID-19 infection by disrupting T helper 17 and regulatory T cell responses, thereby contributing to sustained inflammatory dysfunction and ultimately leading to fatigue.^10^ This inflammatory milieu has been hypothesized to induce an inflammatory feedback loop mediated by a persistent and exaggerated cellular immune response to cytokine release or chronically elevated cytokine release.^1^

These findings have motivated the hypothesis that IL-6 modulating medications, such as tocilizumab and sarilumab, may reduce the risk of Long COVID. Both randomized and observational studies have generally supported that IL-6 receptor antagonists, sarilumab and tocilizumab, may reduce mortality among COVID-19 patients, although further investigation is needed regarding their impact on Long COVID.^11–15^

This study seeks to evaluate the relationship between prescription to IL-6 receptor antagonists (sarilumab or tocilizumab) vs. prescription to other biologic agents (anakinra or baricitinib) and the subsequent risk of severe post-COVID-19 outcomes in a sample of patients with rheumatoid arthritis.

## METHODS

### Sample

This study included electronic health record data for patients in the National Clinical Cohort Collaborative (N3C). N3C offers a rich, high-dimensional data source for a national sample of patients in order to make meaningful inferences regarding the long-term sequelae of COVID-19. N3C includes more than 31 billion rows of data and 8 million COVID-positive patients and 14 million demographically matched controls from 83 data-providing health institutions.^16,17^ N3C identifies patients with acute COVID-19 infection as patients who had either (1) at least one laboratory test with a positive result, (2) at least one “strong positive” diagnostic code in ICD-10 or SNOMED, (3) at least two “weak positive” diagnostic codes in ICD-10 or SNOMED, and selects two demographically-matched controls for each case.^18^ We sampled patients from N3C regardless of past or future COVID-19 status (i.e., including both cases and controls) to avoid inducing bias by restricting on a factor that is an effect of the exposure of interest.^19–24^

### Restriction

We restricted to only include data sites that meet a minimum threshold of reporting for the exposure and outcome of interest (immune modulator prescription and Long COVID) among our target population, defined by more than one standard deviation below the mean reporting rate. These sites have a high risk of (potentially differential) misclassification, and this method is consistent with previous literature.^25,26^

### Temporal Window

We included patients in N3C with a diagnosis of moderate-to-severe rheumatoid arthritis (severity inferred from prescription of a biologic agent) who were prescribed a study drug between December 31, 2021, and January 1, 2023. This ensures that (1) all person-time at risk for Long COVID took place after the release of the Long COVID diagnostic code (ICD code U09.9, “post COVID-19 condition, unspecified,” released October 1, 2021),^27^ (2) COVID-19 infections took place during the Omicron period (supporting consistency in viral and diagnostic trends),^25,28,29^ (3) all patients had equivalent (12 months) outcome monitoring time (i.e., no administrative censoring).

### Inclusion Criteria

We included patients with moderate or severe rheumatoid arthritis (as mild rheumatoid arthritis would not indicate prescription of a biologic agent) who were prescribed one of the following biologic agents, tocilizumab, sarilumab, baricitinib, or anakinra during the study enrollment period and had not been prescribed another study drug in the previous three months. We excluded patients who were prescribed a study drug for severe COVID-19 infection.

### Covariates

We included the following individual-level covariates that may be related to rheumatoid arthritis, provider prescribing decisions, COVID-19 severity, and Long COVID: healthcare utilization rate (described further below), sociodemographic information (sex, age at acute COVID-19 infection, race and ethnicity), comorbidities (bipolar disorder, immunocompromised status, body mass index, diabetes (complicated and/or uncomplicated), chronic lung disease, congestive heart failure, heart failure, acute kidney injury, myocardial infarction, hypertension, asthma, depression, Charlson Comorbidity Index), medication and tobacco usage (tobacco smoking status, systemic corticosteroids), COVID-19 related factors (total number of COVID-19 vaccination and booster doses, indicators of COVID-19 vaccination 0 to 6 months and 6 to 12 months before enrollment), and date of enrollment (prescription).^30,31^ In addition, we adjusted for the following county-level socioeconomic covariates, given the lack of individual-level socioeconomic information in N3C: percent of households with income below the federal poverty line and the social vulnerability index. As healthcare utilization is highly associated with Long COVID diagnosis, we accounted for heterogeneous healthcare utilization in multiple ways. We adjusted for baseline (prior to the exposure of interest, i.e. enrollment) healthcare utilization, defined as the monthly healthcare visitation rate (healthcare visits per month from December 2018 [beginning of N3C observation period] to enrollment). In addition, we considered healthcare utilization during follow-up as a source of informative censoring (i.e., truncation), and we evaluated the counterfactual impact of the exposure of interest, given that all participants were monitored (i.e., had a healthcare interaction) at least once during the 12 months of follow-up.^30,32–34^

### Exposures

Exposures of interest include prescription to tocilizumab or sarilumab (monoclonal antibodies to IL-6), baricitinib (a small molecule inhibitor of JAK2), or anakinra (an IL-1 receptor antagonist), which are FDA-approved immune-modulating biologic agents.^35^ These immune modulators have all received FDA emergency use access or full approval for the treatment of severe COVID-19-related respiratory disease and have been used broadly throughout the pandemic.^36^ Additionally, these medications are FDA-approved for the treatment of rheumatoid arthritis with persistent symptoms despite disease-modifying anti-rheumatic therapy.^35^ As a result, there is a subset of patients with chronic exposure to these targeted immune-modulating therapies who were subsequently infected with SARS-CoV-2. We considered tocilizumab and sarilumab to be the “intervention” drugs, while baricitinib and anakinra were active comparators.

### Outcomes

We evaluated 12-month cumulative incidence of (1) diagnosis of Long COVID based on U09.9 diagnosis code,^27^ (2) probable Long COVID, and (3) mortality as the primary outcomes of interest. The Long COVID computational phenotype is a validated measure that calculates probable Long COVID using individual EHR metrics, and previous studies have used a threshold of 0.9 to indicate probable Long COVID. ^26,37–41^ We evaluated COVID-19 incidence and severe COVID-19 incidence as secondary outcomes. We defined “severe” COVID-19 as any patient who was hospitalized, received invasive ventilation, or died due to COVID-19.

### Analysis Methods

This analysis used an active comparator new user design.^42,43^ First, we applied Super Learner to maximize prediction of the outcome, given treatment and covariates (outcome regression), and model the probability of treatment (prescription of monoclonal antibodies to IL-6, compared to baricitinib and anakinra) given individual covariate status (i.e., treatment mechanism). We included the following nonparametric and nonparametric candidates in our Super Learner library to minimize the risk of model misspecification: generalized linear models (“SL.glm”), GLM net (“SL.glmnet”), and XGBoost (“SL.xgboost”).^44^ Super Learner is well-suited to this data setting, where EHR allows investigators to adjust for high-dimensional covariate data (i.e., many covariates per individual), which would lead to model misspecification via traditional parametric analysis approaches.^22,44,45^ We applied targeted maximum likelihood estimation to analyze the impact of the prescription of monoclonal antibodies to IL-6, compared to baricitinib and anakinra, on the 12-month cumulative incidence of Long COVID among individuals with rheumatoid arthritis.^20,22^ Targeted maximum likelihood estimation is a bias reduction tool that allows us to incorporate causal inference considerations of confounding and temporality to evaluate our causal parameter of interest. Our primary analysis evaluated treatment based on the first drug prescribed during the study period (emulating an intention-to-treat design). We considered death or lack of healthcare utilization (i.e., no healthcare interactions in 12 months after treatment initiation) as informative censoring, and we estimated the counterfactual impact of the intervention under a scenario of universal observation (i.e., at least one healthcare interaction during outcome period and no death [except for analyses of death as the primary outcome of interest]).^34^

### Secondary Analysis

We evaluated the relationship between immune modulator use and severe post-COVID-19 outcomes, restricting to patients with documented COVID-19 prior to enrollment (i.e., prescribed study drug after acute COVID-19) and adjusting for COVID-19 severity (4-point ordinal score). This secondary analysis is analogous to the controlled direct effect of immune-modulating drugs on severe post-COVID outcomes among patients with COVID-19, excluding the indirect pathway mediated through Long COVID incidence and severity.^46^ In other words, this secondary analysis evaluated whether the impact of immune-modulating drugs on severe post-COVID-19 outcomes is mediated by COVID-19 incidence and severity, or whether there is a direct pathway from these drugs to Long COVID or mortality.

## RESULTS

**Table 1.**
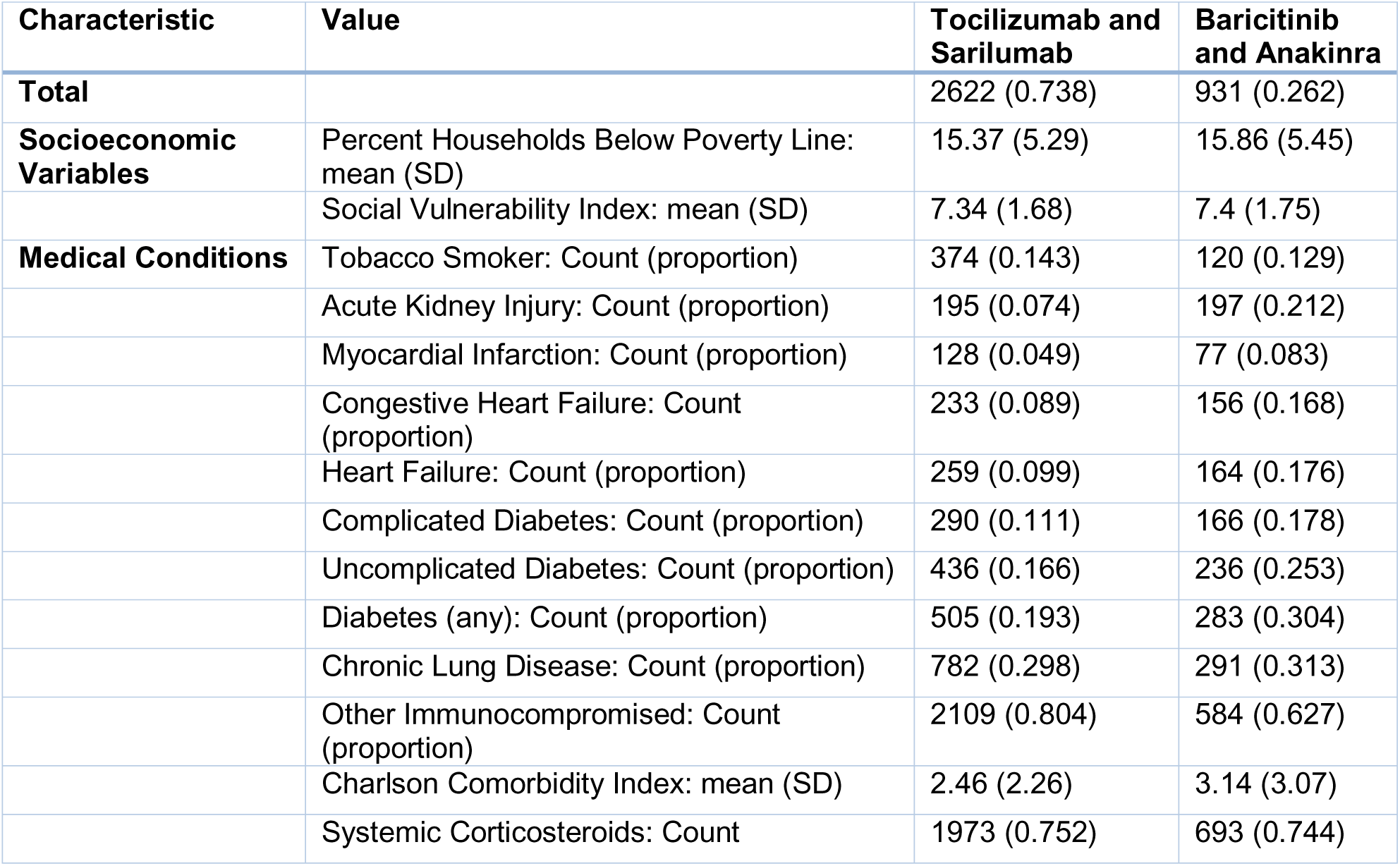
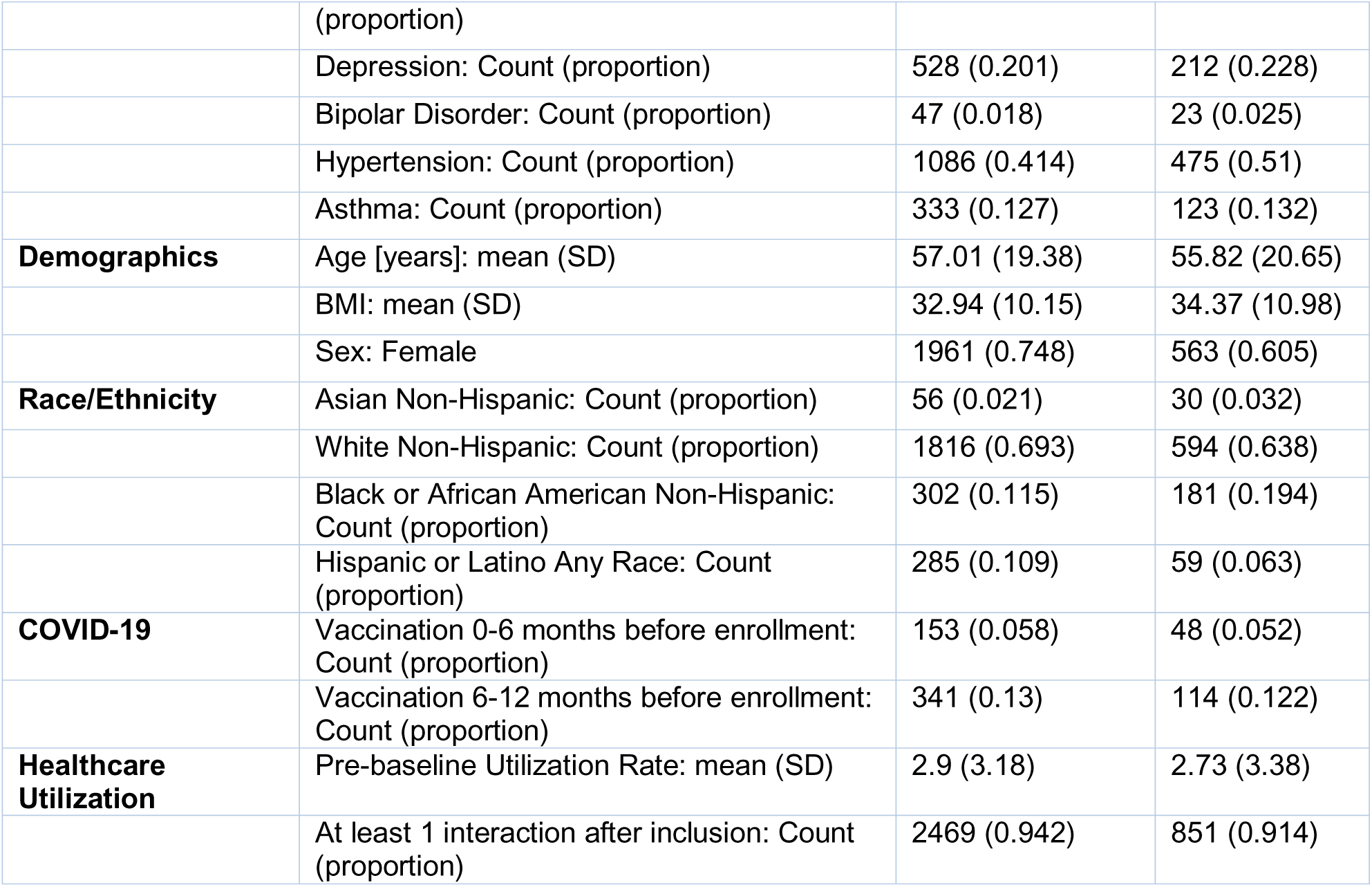
Patients with moderate to severe rheumatoid arthritis who were prescribed tocilizumab, sarilumab (treatment), anakinra, or baricitinib (control) in 2022 in the National Clinical Cohort Collaborative.

| Characteristic | Value | Tocilizumab and Sarilumab | Baricitinib and Anakinra |
| --- | --- | --- | --- |
| **Total** |  | 2622 (0.738) | 931 (0.262) |
| **Socioeconomic Variables** | Percent Households Below Poverty Line: mean (SD) | 15.37 (5.29) | 15.86 (5.45) |
|  | Social Vulnerability Index: mean (SD) | 7.34 (1.68) | 7.4 (1.75) |
| **Medical Conditions** | Tobacco Smoker: Count (proportion) | 374 (0.143) | 120 (0.129) |
|  | Acute Kidney Injury: Count (proportion) | 195 (0.074) | 197 (0.212) |
|  | Myocardial Infarction: Count (proportion) | 128 (0.049) | 77 (0.083) |
|  | Congestive Heart Failure: Count (proportion) | 233 (0.089) | 156 (0.168) |
|  | Heart Failure: Count (proportion) | 259 (0.099) | 164 (0.176) |
|  | Complicated Diabetes: Count (proportion) | 290 (0.111) | 166 (0.178) |
|  | Uncomplicated Diabetes: Count (proportion) | 436 (0.166) | 236 (0.253) |
|  | Diabetes (any): Count (proportion) | 505 (0.193) | 283 (0.304) |
|  | Chronic Lung Disease: Count (proportion) | 782 (0.298) | 291 (0.313) |
|  | Other Immunocompromised: Count (proportion) | 2109 (0.804) | 584 (0.627) |
|  | Charlson Comorbidity Index: mean (SD) | 2.46 (2.26) | 3.14 (3.07) |
|  | Systemic Corticosteroids: Count (proportion) | 1973 (0.752) | 693 (0.744) |
|  | Depression: Count (proportion) | 528 (0.201) | 212 (0.228) |
|  | Bipolar Disorder: Count (proportion) | 47 (0.018) | 23 (0.025) |
|  | Hypertension: Count (proportion) | 1086 (0.414) | 475 (0.51) |
|  | Asthma: Count (proportion) | 333 (0.127) | 123 (0.132) |
| **Demographics** | Age [years]: mean (SD) | 57.01 (19.38) | 55.82 (20.65) |
|  | BMI: mean (SD) | 32.94 (10.15) | 34.37 (10.98) |
|  | Sex: Female | 1961 (0.748) | 563 (0.605) |
| **Race/Ethnicity** | Asian Non-Hispanic: Count (proportion) | 56 (0.021) | 30 (0.032) |
|  | White Non-Hispanic: Count (proportion) | 1816 (0.693) | 594 (0.638) |
|  | Black or African American Non-Hispanic: Count (proportion) | 302 (0.115) | 181 (0.194) |
|  | Hispanic or Latino Any Race: Count (proportion) | 285 (0.109) | 59 (0.063) |
| **COVID-19** | Vaccination 0-6 months before enrollment: Count (proportion) | 153 (0.058) | 48 (0.052) |
|  | Vaccination 6-12 months before enrollment: Count (proportion) | 341 (0.13) | 114 (0.122) |
| **Healthcare Utilization** | Pre-baseline Utilization Rate: mean (SD) | 2.9 (3.18) | 2.73 (3.38) |
|  | At least 1 interaction after inclusion: Count (proportion) | 2469 (0.942) | 851 (0.914) |

We analyzed electronic health record data from a sample of 3,553 patients with rheumatoid arthritis who were prescribed tocilizumab, sarilumab, anakinra, or baricitinib in 2022. In our sample, 2,622 patients were taking treatment drugs (tocilizumab or sarilumab), while 931 patients were taking control drugs (anakinra or baricitinib). The average age of treatment patients was 57 years, while the average age of comparator patients was 56 years. Most patients were White non-Hispanic (69% in the treatment group, 64% in the comparator group). Treatment patients had an average Charlson Comorbidity Index of 2.5, compared to 3.1 for comparator patients.

**Figure 1.**
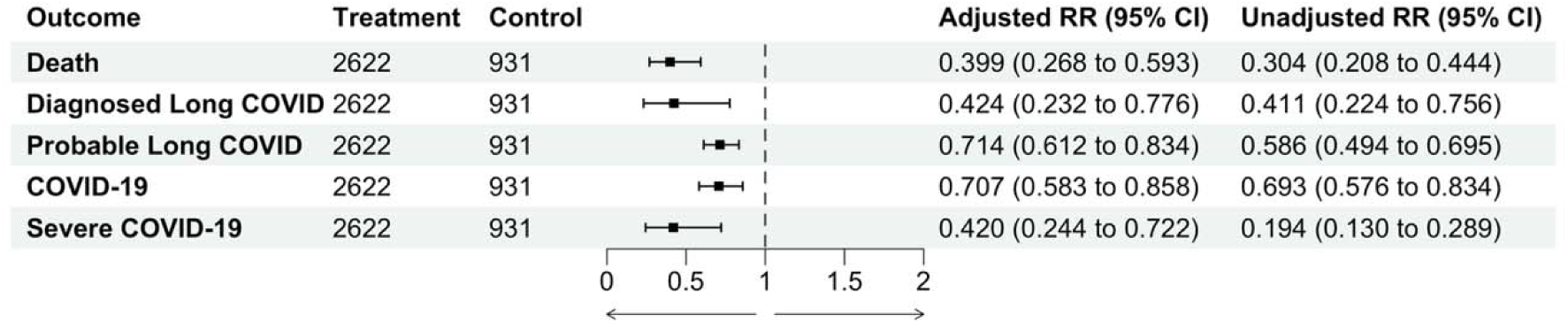
Relationship between IL-6 modulating drugs (tocilizumab or sarilumab) vs. other biologic agents (anakinra or baricitinib) and 12-month cumulative incidence of post-COVID-19 and acute COVID-19 outcomes, among patients with moderate to severe rheumatoid arthritis prescribed a study drug in 2022.

We found that prescription to IL-6 modulating drugs, compared to prescription to other biologic agents, was associated with a lower 12-month cumulative risk of mortality (adjusted relative risk (aRR) 0.40, 95% CI 0.27, 0.59), diagnosed Long COVID (aRR 0.42, 95% CI 0.23, 0.78), and probable Long COVID (aRR 0.71, 95% CI 0.61, 0.83). We found that prescription to IL-6 modulating drugs was associated with a lower 12-month cumulative incidence of COVID-19 (aRR 0.71, 95% CI 0.58, 0.86) or severe COVID-19 (aRR 0.42, 95% CI 0.24, 0.72), compared with prescription of other biologic agents.

In our secondary analysis, restricting our sample to only include patients who were prescribed a study drug after acute COVID-19, we did not find a significant controlled direct effect of IL-6 modulating drugs, compared with other biologic agents, on the cumulative risk of mortality (controlled direct effect adjusted relative risk (aRR_CDE_) 0.95, 95% CI 0.39, 2.39), diagnosed Long COVID (aRR_CDE_ 0.53, 95% CI 0.20, 1.41), or probable Long COVID (aRR_CDE_ 0.90, 95% CI 0.75, 1.06), among COVID-19-positive patients (Supplemental Material 1).

## DISCUSSION

We found evidence that prescription to IL-6 modulating drugs (sarilumab or tocilizumab), compared to other biologic agents, was associated with a lower risk of multiple severe post-COVID-19 outcomes (death, Long COVID diagnosis, and probable Long COVID) in a cohort of patients with rheumatoid arthritis. This provides support for the benefit of IL-6 modulating drugs for rheumatoid arthritis patients during the COVID-19 pandemic, particularly patients who may be vulnerable to severe COVID-19. Furthermore, these findings support the role of IL-6 in influencing COVID-19 severity and the development of severe long-term sequelae of COVID-19, such as death or Long COVID.

IL-6 signaling plays a crucial role in mediating systemic inflammation and endothelial function, which are key mechanisms of acute COVID-19 and post-acute sequelae of COVID-19.^47–49^ IL-6 receptor antagonists may interrupt this pathway during acute infection and prevent the long-term consequences of severe infection. In addition, previous studies have consistently found elevated IL-6 levels among Long COVID patients and have hypothesized that IL-6 may mediate long-term neurological symptoms of COVID-19 infection by disrupting T helper 17 and regulatory T cell responses, contributing to sustained inflammatory dysfunction and ultimately leading to fatigue.^10^

We did not find a significant, controlled direct effect of IL-6 modulating drugs, compared with other biologic agents, on post-COVID-19 outcomes among COVID-19-positive individuals, indicating that the protective effects of IL-6 modulating drugs are largely mediated by reduced risk of COVID-19 and reduced COVID-19 severity. In our secondary analysis, we evaluated a subset of our total sample in which the drug start date (new IL-6 medications or comparator) was after acute COVID-19 incidence. While we found a significant protective effect of IL-6 medications on severe post-COVID-19 outcomes in patients who were prescribed IL-6 medications before COVID-19, we found no protective effect in patients prescribed IL-6 medications after COVID-19. Therefore, our findings support that only IL-6 modulating drugs delivered before acute COVID-19, not during or after acute COVID-19, may protect against long-term sequelae of COVID-19.

### Strengths, limitations, and areas for future study

The EHR data source was a considerable strength of this study, as it contained a large sample size of patients and a wide range of covariate information for each patient.^50^ The analysis approach, leveraging Super Learner and targeted maximum likelihood estimation, was a second strength of this study, as it enabled semiparametric evaluation of this research question using a doubly-robust estimation method, minimizing bias and confounding.^19–22,51^

Our findings regarding the protective relationship between IL-6 modulating drugs and acute COVID-19 severity are consistent with previous studies. N3C’s sampling methodology includes a test-negative design of COVID-19 cases and sociodemographically matched controls who seek COVID-19 testing at a testing facility.^16,17^ Our finding that IL-6 modulating drugs were associated with reduced COVID-19 incidence may be a consequence of this sampling methodology and reduced COVID-19 severity in IL-6 patients, as patients who are asymptomatic or have mild COVID-19 may not seek SARS-CoV-2 testing at a healthcare facility. Therefore, we do not assume a causal relationship between IL-6 RA medications and COVID-19 incidence.

The generalizability of N3C is a limitation, as it oversamples patients with high healthcare-seeking behavior, leading to an overrepresentation of patients with multiple comorbidities, who are white, and who are older.^38,40,41^ While our study specifically seeks to evaluate the relationship between IL-6 and Long COVID, the interdependence of inflammation and cytokines complicates these pathways.^3,7,47^ Similarly, the use of biologic agents has myriad effects across immune pathways. There is also a possibility of residual confounding by indication, as patients with rheumatoid arthritis who were prescribed tocilizumab, sarilumab, anakinra, or baricitinib may reflect unique populations. While we sought to account for this baseline imbalance through covariate adjustment and doubly robust estimation, the possibility of residual confounding remains a concern. The low sensitivity of Long COVID diagnosis is a limitation, as Long COVID is rarely diagnosed and documented in EHR due to the wide range of phenotypic manifestations and few treatment options for Long COVID patients.^38,40,52^ We sought to overcome this potential limitation by including probable Long COVID as an additional outcome, which has greater sensitivity than a Long COVID diagnosis but may have less clinical utility. Finally, biomarker data are limited in N3C, which precludes a direct measurement of IL-6 biomarkers. Future studies should include direct biomarker evaluation to determine the mechanistic role of IL-6 in the development of severe post-COVID-19 outcomes.

## CONCLUSIONS

Among patients with rheumatoid arthritis, we found that prescription of IL-6 modulating medications was associated with reduced risk of long-term sequelae of COVID-19, which may be mediated through protection against COVID-19 incidence and severity. This supports the utility of IL-6 modulating drugs for rheumatoid arthritis patients who are at risk of severe COVID-19 and provides indirect support for IL-6 as a mechanistic biomarker of long-term sequelae of COVID-19.

## Data Availability

All analytic code and data are available in the N3C Enclave by request. Access to the N3C Data Enclave is managed by NCATS (https://ncats.nih.gov/research/research-activities/n3c/resources/data-access). Interested researchers must first complete a data use agreement, and next a data use request, in order to access the N3C Data Enclave. Once access is granted, the N3C data use committee must review and approve all use of data and the publication committee must approve all publications involving N3C data.

## DECLARATIONS

## Ethics approval and consent to participate

This study was approved by the UC Berkeley Office for Protection of Human Subjects (2022-01-14980). The N3C data transfer to NCATS is performed under a Johns Hopkins University Reliance Protocol # IRB00249128 or individual site agreements with NIH. N3C received a waiver of consent from the NIH Institutional Review board and allows the secondary analysis of these data without additional consent.

## Consent to publish

The authors consent to the publication of this manuscript in its entirety.

Availability of data and materials: All analytic code and data are available in the N3C Enclave by request. Access to the N3C Data Enclave is managed by NCATS (https://ncats.nih.gov/research/research-activities/n3c/resources/data-access). Interested researchers must first complete a data use agreement, and next a data use request, in order to access the N3C Data Enclave. Once access is granted, the N3C data use committee must review and approve all use of data and the publication committee must approve all publications involving N3C data.

## Funding

This research was financially supported by the National Institute for Allergy and Infectious Diseases (1K01AI182501-01 to Zachary Butzin-Dozier). Individual authors were supported by the following funding sources: NIMH R01131542 (PI Rena C. Patel), Jerrod Anzalone is supported by the National Institute of General Medical Sciences, U54 GM115458, which funds the Great Plains IDeA-CTR Network. The content is solely the responsibility of the authors and does not necessarily represent the official views of the NIH.

## N3C Attribution

The analyses described in this manuscript were conducted with data or tools accessed through the NCATS N3C Data Enclave https://covid.cd2h.org and N3C Attribution & Publication Policy v 1.2-2020-08-25b supported by NCATS U24 TR002306, Axle Informatics Subcontract: NCATS-P00438-B, the Bill & Melinda Gates Foundation: OPP1165144, and the National Institutes of General Medical Sciences: U54GM115458 and 5U54GM104942-04. This research was possible because of the patients whose information is included within the data and the organizations (https://ncats.nih.gov/n3c/resources/data-contribution/data-transfer-agreement-signatories) and scientists who have contributed to the ongoing development of this community resource [https://doi.org/10.1093/jamia/ocaa196].

## Disclaimer

The N3C Publication committee confirmed that this manuscript (MSID:2738.642) complies with N3C data use and attribution policies; however, the authors are solely responsible for its content, which does not necessarily represent the official views of the National Institutes of Health or the N3C program.

## IRB

The N3C data transfer to NCATS is performed under a Johns Hopkins University Reliance Protocol # IRB00249128 or individual site agreements with NIH. The N3C Data Enclave is managed under the authority of the NIH; information can be found at https://ncats.nih.gov/n3c/resources.

This research project was approved by the University of California, Berkeley Committee for the Protection of Human Subjects (CPHS protocol number 2022-01-14980). This approval is issued under University of California, Berkeley Federalwide Assurance #00006252.

## Individual Acknowledgements For Core Contributors

We gratefully acknowledge the following core contributors to N3C:

Adam B. Wilcox, Adam M. Lee, Alexis Graves, Alfred (Jerrod) Anzalone, Amin Manna, Amit Saha, Amy Olex, Andrea Zhou, Andrew E. Williams, Andrew Southerland, Andrew T. Girvin, Anita Walden, Anjali A. Sharathkumar, Benjamin Amor, Benjamin Bates, Brian Hendricks, Brijesh Patel, Caleb Alexander, Carolyn Bramante, Cavin Ward-Caviness, Charisse Madlock-Brown, Christine Suver, Christopher Chute, Christopher Dillon, Chunlei Wu, Clare Schmitt, Cliff Takemoto, Dan Housman, Davera Gabriel, David A. Eichmann, Diego Mazzotti, Don Brown, Eilis Boudreau, Elaine Hill, Elizabeth Zampino, Emily Carlson Marti, Emily R. Pfaff, Evan French, Farrukh M Koraishy, Federico Mariona, Fred Prior, George Sokos, Greg Martin, Harold Lehmann, Heidi Spratt, Hemalkumar Mehta, Hongfang Liu, Hythem Sidky, J.W. Awori Hayanga, Jami Pincavitch, Jaylyn Clark, Jeremy Richard Harper, Jessica Islam, Jin Ge, Joel Gagnier, Joel H. Saltz, Joel Saltz, Johanna Loomba, John Buse, Jomol Mathew, Joni L. Rutter, Julie A. McMurry, Justin Guinney, Justin Starren, Karen Crowley, Katie Rebecca Bradwell, Kellie M. Walters, Ken Wilkins, Kenneth R. Gersing, Kenrick Dwain Cato, Kimberly Murray, Kristin Kostka, Lavance Northington, Lee Allan Pyles, Leonie Misquitta, Lesley Cottrell, Lili Portilla, Mariam Deacy, Mark M. Bissell, Marshall Clark, Mary Emmett, Mary Morrison Saltz, Matvey B. Palchuk, Melissa A. Haendel, Meredith Adams, Meredith Temple-O’Connor, Michael G. Kurilla, Michele Morris, Nabeel Qureshi, Nasia Safdar, Nicole Garbarini, Noha Sharafeldin, Ofer Sadan, Patricia A. Francis, Penny Wung Burgoon, Peter Robinson, Philip R.O. Payne, Rafael Fuentes, Randeep Jawa, Rebecca Erwin-Cohen, Rena Patel, Richard A. Moffitt, Richard L. Zhu, Rishi Kamaleswaran, Robert Hurley, Robert T. Miller, Saiju Pyarajan, Sam G. Michael, Samuel Bozzette, Sandeep Mallipattu, Satyanarayana Vedula, Scott Chapman, Shawn T. O’Neil, Soko Setoguchi, Stephanie S. Hong, Steve Johnson, Tellen D. Bennett, Tiffany Callahan, Umit Topaloglu, Usman Sheikh, Valery Gordon, Vignesh Subbian, Warren A. Kibbe, Wenndy Hernandez, Will Beasley, Will Cooper, William Hillegass, Xiaohan Tanner Zhang. Details of contributions available at covid.cd2h.org/core-contributors

## Data Partners with Released Data

The following institutions whose data is released or pending:

Available: Advocate Health Care Network — UL1TR002389: The Institute for Translational Medicine (ITM) • Boston University Medical Campus — UL1TR001430: Boston University Clinical and Translational Science Institute • Brown University — U54GM115677: Advance Clinical Translational Research (Advance-CTR) • Carilion Clinic — UL1TR003015: iTHRIV Integrated Translational health Research Institute of Virginia • Charleston Area Medical Center — U54GM104942: West Virginia Clinical and Translational Science Institute (WVCTSI) • Children’s Hospital Colorado — UL1TR002535: Colorado Clinical and Translational Sciences Institute • Columbia University Irving Medical Center — UL1TR001873: Irving Institute for Clinical and Translational Research • Duke University — UL1TR002553: Duke Clinical and Translational Science Institute • George Washington Children’s Research Institute — UL1TR001876: Clinical and Translational Science Institute at Children’s National (CTSA-CN) • George Washington University — UL1TR001876: Clinical and Translational Science Institute at Children’s National (CTSA-CN) • Indiana University School of Medicine — UL1TR002529: Indiana Clinical and Translational Science Institute • Johns Hopkins University — UL1TR003098: Johns Hopkins Institute for Clinical and Translational Research • Loyola Medicine — Loyola University Medical Center • Loyola University Medical Center — UL1TR002389: The Institute for Translational Medicine (ITM) • Maine Medical Center — U54GM115516: Northern New England Clinical & Translational Research (NNE-CTR) Network • Massachusetts General Brigham — UL1TR002541: Harvard Catalyst • Mayo Clinic Rochester — UL1TR002377: Mayo Clinic Center for Clinical and Translational Science (CCaTS) • Medical University of South Carolina — UL1TR001450: South Carolina Clinical & Translational Research Institute (SCTR) • Montefiore Medical Center — UL1TR002556: Institute for Clinical and Translational Research at Einstein and Montefiore • Nemours U54GM104941: Delaware CTR ACCEL Program • NorthShore University HealthSystem — UL1TR002389: The Institute for Translational Medicine (ITM) • Northwestern University at Chicago — UL1TR001422: Northwestern University Clinical and Translational Science Institute (NUCATS) • OCHIN — INV-018455: Bill and Melinda Gates Foundation grant to Sage Bionetworks • Oregon Health & Science University — UL1TR002369: Oregon Clinical and Translational Research Institute • Penn State Health Milton S. Hershey Medical Center — UL1TR002014: Penn State Clinical and Translational Science Institute • Rush University Medical Center — UL1TR002389: The Institute for Translational Medicine (ITM) • Rutgers, The State University of New Jersey UL1TR003017: New Jersey Alliance for Clinical and Translational Science • Stony Brook University — U24TR002306 • The Ohio State University — UL1TR002733: Center for Clinical and Translational Science • The State University of New York at Buffalo — UL1TR001412: Clinical and Translational Science Institute • The University of Chicago — UL1TR002389: The Institute for Translational Medicine (ITM) • The University of Iowa — UL1TR002537: Institute for Clinical and Translational Science • The University of Miami Leonard M. Miller School of Medicine — UL1TR002736: University of Miami Clinical and Translational Science Institute • The University of Michigan at Ann Arbor — UL1TR002240: Michigan Institute for Clinical and Health Research • The University of Texas Health Science Center at Houston — UL1TR003167: Center for Clinical and Translational Sciences (CCTS) • The University of Texas Medical Branch at Galveston — UL1TR001439: The Institute for Translational Sciences • The University of Utah — UL1TR002538: Uhealth Center for Clinical and Translational Science • Tufts Medical Center — UL1TR002544: Tufts Clinical and Translational Science Institute • Tulane University — UL1TR003096: Center for Clinical and Translational Science • University Medical Center New Orleans — U54GM104940: Louisiana Clinical and Translational Science (LA CaTS) Center • University of Alabama at Birmingham — UL1TR003096: Center for Clinical and Translational Science • University of Arkansas for Medical Sciences — UL1TR003107: UAMS Translational Research Institute • University of Cincinnati — UL1TR001425: Center for Clinical and Translational Science and Training • University of Colorado Denver, Anschutz Medical Campus — UL1TR002535: Colorado Clinical and Translational Sciences Institute • University of Illinois at Chicago — UL1TR002003: UIC Center for Clinical and Translational Science • University of Kansas Medical Center — UL1TR002366: Frontiers: University of Kansas Clinical and Translational Science Institute • University of Kentucky — UL1TR001998: UK Center for Clinical and Translational Science • University of Massachusetts Medical School Worcester — UL1TR001453: The UMass Center for Clinical and Translational Science (UMCCTS) • University of Minnesota — UL1TR002494: Clinical and Translational Science Institute • University of Mississippi Medical Center — U54GM115428: Mississippi Center for Clinical and Translational Research (CCTR) • University of Nebraska Medical Center — U54GM115458: Great Plains IDeA-Clinical & Translational Research • University of North Carolina at Chapel Hill — UL1TR002489: North Carolina Translational and Clinical Science Institute • University of Oklahoma Health Sciences Center — U54GM104938: Oklahoma Clinical and Translational Science Institute (OCTSI) • University of Rochester — UL1TR002001: UR Clinical & Translational Science Institute • University of Southern California — UL1TR001855: The Southern California Clinical and Translational Science Institute (SC CTSI) • University of Vermont — U54GM115516: Northern New England Clinical & Translational Research (NNE-CTR) Network • University of Virginia — UL1TR003015: iTHRIV Integrated Translational health Research Institute of Virginia • University of Washington — UL1TR002319: Institute of Translational Health Sciences • University of Wisconsin-Madison — UL1TR002373: UW Institute for Clinical and Translational Research • Vanderbilt University Medical Center — UL1TR002243: Vanderbilt Institute for Clinical and Translational Research • Virginia Commonwealth University — UL1TR002649: C. Kenneth and Dianne Wright Center for Clinical and Translational Research • Wake Forest University Health Sciences — UL1TR001420: Wake Forest Clinical and Translational Science Institute • Washington University in St. Louis — UL1TR002345: Institute of Clinical and Translational Sciences • Weill Medical College of Cornell University — UL1TR002384: Weill Cornell Medicine Clinical and Translational Science Center • West Virginia University — U54GM104942: West Virginia Clinical and Translational Science Institute (WVCTSI)

## Submitted

Icahn School of Medicine at Mount Sinai — UL1TR001433: ConduITS Institute for Translational Sciences • The University of Texas Health Science Center at Tyler — UL1TR003167: Center for Clinical and Translational Sciences (CCTS) • University of California, Davis — UL1TR001860: UCDavis Health Clinical and Translational Science Center • University of California, Irvine — UL1TR001414: The UC Irvine Institute for Clinical and Translational Science (ICTS) • University of California, Los Angeles — UL1TR001881: UCLA Clinical Translational Science Institute • University of California, San Diego — UL1TR001442: Altman Clinical and Translational Research Institute • University of California, San Francisco — UL1TR001872: UCSF Clinical and Translational Science Institute

## Pending

Arkansas Children’s Hospital — UL1TR003107: UAMS Translational Research Institute • Baylor College of Medicine — None (Voluntary) • Children’s Hospital of Philadelphia — UL1TR001878: Institute for Translational Medicine and Therapeutics • Cincinnati Children’s Hospital Medical Center — UL1TR001425: Center for Clinical and Translational Science and Training • Emory University — UL1TR002378: Georgia Clinical and Translational Science Alliance • HonorHealth — None (Voluntary) • Loyola University Chicago — UL1TR002389: The Institute for Translational Medicine (ITM) • Medical College of Wisconsin — UL1TR001436: Clinical and Translational Science Institute of Southeast Wisconsin • MedStar Health Research Institute — UL1TR001409: The Georgetown-Howard Universities Center for Clinical and Translational Science (GHUCCTS) • MetroHealth — None (Voluntary) • Montana State University — U54GM115371: American Indian/Alaska Native CTR • NYU Langone Medical Center — UL1TR001445: Langone Health’s Clinical and Translational Science Institute • Ochsner Medical Center — U54GM104940: Louisiana Clinical and Translational Science (LA CaTS) Center • Regenstrief Institute — UL1TR002529: Indiana Clinical and Translational Science Institute • Sanford Research — None (Voluntary) • Stanford University — UL1TR003142: Spectrum: The Stanford Center for Clinical and Translational Research and Education • The Rockefeller University — UL1TR001866: Center for Clinical and Translational Science • The Scripps Research Institute — UL1TR002550: Scripps Research Translational Institute • University of Florida — UL1TR001427: UF Clinical and Translational Science Institute • University of New Mexico Health Sciences Center — UL1TR001449: University of New Mexico Clinical and Translational Science Center • University of Texas Health Science Center at San Antonio — UL1TR002645: Institute for Integration of Medicine and Science • Yale New Haven Hospital — UL1TR001863: Yale Center for Clinical Investigation

## Authors statement

Authorship was determined using ICMJE recommendations.

ZB and BS: Generated research question,

ZB: drafted manuscript, managed project timeline, and coordinated analysis.

MK, YJ, LW, AJA, EH, RCP, RW, CB, and BS: Provided oversight on study design and analysis plan, supported analysis, reviewed manuscript, and provided feedback.

## Inclusion and ethics statement

All co-authors and collaborators included in this manuscript have fulfilled the criteria for authorship.

