## Supplemental Material 1 for "IL-6 Receptor Antagonists and Severe Post-COVID-19 Outcomes: An Emulated Target Trial"

**SUPPLEMENTAL MATERIALS**

Supplemental Material 1. Relationships between IL-6 modulating drugs (tocilizumab or sarilumab) vs. other biologic agents (anakinra or baricitinib) and the 12-month cumulative incidence of post-COVID-19 outcomes among type 2 diabetes mellitus patients prescribed a study drug in 2022 with documented acute COVID-19 before the drug start date.
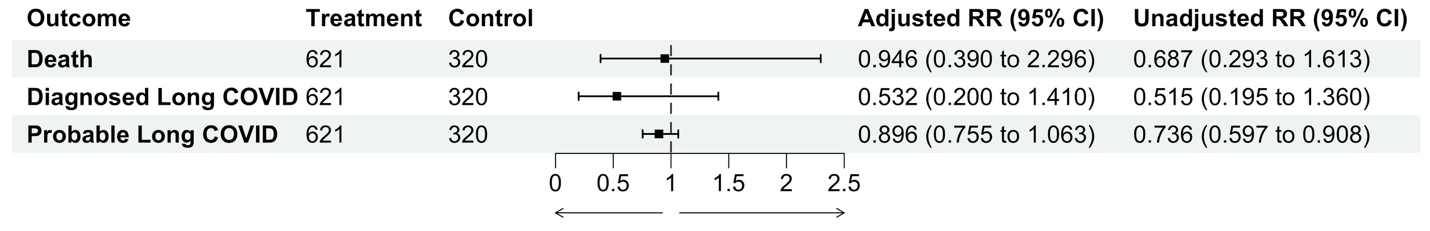
